# The impact of COVID-19 on Long Term Care Facilities (LTCFs) of an Italian Province: a cohort study and a retrospective analysis of observed vs. expected mortality

**DOI:** 10.1101/2020.10.21.20216705

**Authors:** Andrea Tramarin, Nicola Gennaro, Giancarlo Dal Grande, Luciana Bragagnolo, Maria Rosa Carta, Davide Giavarina, Michela Pascarella, Mario Rassu, Antonio Matteazzi, Giampaolo Stopazzolo

## Abstract

In Italy, as in other countries, Long Term Care Facilities (LTCFs) have seen a disproportionally high number of deaths during the COVID-19 pandemic. The Veneto region was one of the first areas of the country where the virus spread rapidly particularly in the LTCFs. As it became evident that LTCs were the epicenter of the pandemic, health authorities of the Vicenza province adopted a plan, which included an epidemiological investigation in a case study facility (CSF) and a retrospective analysis to estimate the impact of COVID-19 in terms of mortality. Combining retrospective data and a prospective cohort study in the CSF we provided a tentative estimate of the impact of COVID-19 on LTCFs. We found an age-gradient in all variables explored. An observed mortality higher 60% than 2019 was found in those LTCF reporting COVID-19 cases. Our findings suggest the need to adopt and maintain strict mitigation measures in LTCFs in the future dynamics of the epidemic.

## Introduction

On December 12, 2020, 27 cases of pneumonia of unknown causes were reported in Wuhan, Hubei Province, China. With the evolving pandemic, COVID-19 spread rapidly from China around the entire world. Italy was the first European nation to be affected by COVID-19. On January 31, two Chinese tourists resulted positive at SARS-2 swab test in Rome. An outbreak of 16 confirmed cases, never been travelling from and to China, were then reported in the Veneto and in the Lombardy Regions. The number of cases rose rapidly with a geometric progression in the two Regions (Veneto and Lombardy) and through all the country. In response to the growing pandemic of COVID-19, the Italian government imposed a national quarantine, restricting the movement of the population except for necessities such as work and health circumstances. On May 31, in the national surveillance system, 233,515 confirmed total cases and 33,530 deaths have been reported [1]. Preliminary studies found that, at a community-level, COVID-19 had a rapid spread and high morbidity and mortality among older adults in Long Term Care Facilities (LTCFs) [2]. A retrospective analysis of individual cases data from China, and elsewhere, showed a strong age gradient in the case fatality ratio [3]. Cardiovascular diseases, hypertension, diabetes mellitus were the comorbidities most frequently associated with COVID-19. All these comorbidities are common among dependent elderly housed in institution and many elderly died by the association between their original comorbidities and the novel virus [4].

In the initial emergency, data from LTCFs not only stressed the vulnerability of its patients and residents and this led to national headlines. Major Italian newspapers reported figures and accounts of incredibly high numbers of deaths in residential care settings denouncing lack of guidelines, medical procedures, testing for COVID-19 and supply of PPE. A dedicated survey has been done by the National Institute of Health in the month of April. The responding LTCFs reported a mortality of 8.4% in the month of March. Among the 3859 total deaths, only 133 were officially classified as COVID-19 after appropriate testing though, 1,310 had flu and COVID-19 related symptoms. The National Institute of Health affirms that these two numbers should be analyzed jointly accounting for the 37.4% of the deaths of the period as COVID-19 related [5]. However, the real impact in terms of mortality in LTCFs is still unknown in Italy.

The term LTCF encompasses a diverse range of healthcare settings including nursing homes, rehabilitation centers, long-term care hospitals, psychiatric care facilities and facilities for people with intellectual disabilities. Although people of all ages may reside in these facilities, the majority of residents are elderly.

This paper aims to illustrate the multiple aspects of an outbreak of COVID-19 in a community of aged individuals and to describe the role of ageing in the interaction between the virus and the host. It might also be considered as a contribution to the current debate on global or selective lockdown assigning to LTCFs, and to aged people, the highest priority in the future facing of the pandemic.

## Background

The Vicenza Province is one of the seven Provinces of the Veneto Region situated in the North East of Italy. It has 867.314 inhabitants. In the AULSS 8, health district there are 34 LTCFs hosting a total 3664 residents. The first COVID-19 case occurred in a LTCF of the Vicenza Province on March 19. After four days of an unexplained fever and worsening respiratory status, a resident of a LTCF was transferred to a local hospital. Two days after, she resulted positive to COVID-19. After the news of the index case, all residents of the facility underwent promptly to a COVID-19 swab tests and the vast majority of them (82%) were positives

After the case-index notification, strict infection control measures have been adopted (i.e. isolation of patients, provision of PPE, disinfection, closure to new admission and visitors, educational support, etc.). In the meanwhile, other sporadic cases started to be notified in other LTCF and local health authorities decided to conduct an in-depth epidemiological investigation in this case facility (CF). According to national headlines which have been reporting a disproportionate high mortality in this care setting a specific mortality registry in all other facilities was adopted.

## Methods

### Phase one: the in-depth prospective epidemiological investigation

An epidemiological investigation was conducted in the CF according to CDC recommendations [6]. The case definition assumed was the positivity to a COVID-19. The rapid response from the laboratory made not necessary the adoption of criteria to define probable or possible cases. Due to the high sensitivity of the molecular tests, the definition of false positives was also not considered necessary. To exclude false negative cases a second round of swabs was repeated after a week in all negative subjects.

The staging WHO definition of illness severity was assumed [7]. All residents have been monitored daily for symptoms and checked by measuring temperature, oxygen saturation via pulse oximetry and respiratory rate at bed site. Asymptomatic were those residents with the absence of signs and symptoms. Mild disease was defined by symptoms without evidence of viral pneumonia or hypoxia; moderate disease was defined as the presence of cough, fever, with sign of pneumonia. severe disease was stated by the presence of fever, cough, plus respiratory rate of > 30 breaths/min or SpO2 < 90% on room air [8].

Recovery was defined by two consecutives negative PCR-RT in a nasopharyngeal swab test which were scheduled at the fourth week after positivity. For a better definition of recovery, a blood drawn was done for measuring the antibodies titer at the fourth week and it was repeated also after 16 weeks.

According to the Italian Society of Geriatrics and Gerontology, all residents were categorized into three groups: 65-74 years-old; 75-84 years-old; 85-99 years-old and centenaries (above 100 years old) [9]. Comorbidity was obtained from the electronic clinical records and appraised selecting those residents affected by less than 2 and those with more than 2 chronic pathologies.

In consideration of the peculiarity of the population under investigation, disability has been considered as a potential risk factor and the Barthel scale was used for its evaluation. The Barthel scale is composed by ten indicators that may be grouped into two main ranks: 1. dependency (i.e. feeding, personal toileting/bathing, dressing/undressing, getting on/off a toilet, controlling bladder/bowel); 2.mobility (moving/returning from wheelchair to bed, walking on level surface or propelling a wheelchair if unable to walk and ascending/descending stairs [10].

The detection of SARS-CoV-2 nucleic acids was performed on Cobas 6800 TR-PCR System (Roche Diagnostic GmbH, Mannheim, Germany). A cycle threshold value (Ct-value) ≤38 was defined as a positive test result and a Ct-value of >40 or no amplification curve was defined as negative. A median value of 24 of Ct was assumed to define high or ow viral load The method to measure SARS-CoV-specific IgM and IgG titer was a chemiluminescent (CLIA) assay (Maglumi 800, SNIBE). Antibodies used in these assays are directed against both CoV-S (spike) and CoV-N (nucleocapside)..

### Phase two: the assessment of COVID-19 on mortality

To assess the impact on mortality by COVID-19, a retrospective cohort study has been carried out in all those LTCFs of the Province with a COVID-19 case notification. Mortality has been evaluated considering the time period from 2020 December 1 to June 15. We used three basic measures: the attack rate, the case fatality rate and the standardized mortality ratio. The attack rate is the percentage of the population that contracts the disease in an at risk population during a specified time interval. The Case Fatality Rate (CFR) is a measure obtained dividing the number of deaths from COVID-19 by the number of cases of COVID-19. The excess of mortality in the COVID-19 LTCFs approach relies on the Standardized Mortality Ratios (SMRs). It is defined as the ratio of observed deaths in the study group compared to expected deaths in the study population. To calculate SMRs, we considered first the number of residents from January 1 to June 15 on 2018, 2019 and 2020. Secondly, we calculated and compared the ratio between age- and gender-specific deaths observed with those expected in these time periods. Data were obtained from centralized demographic data.

#### Statistics

The chi-square test was used to assess differences in demographics and clinical characteristics according to attack rate, CFR and the time to negative conversion of viral RNA. The trend in the age group was evaluated through a test for trend estimated using a logistic regression model. Data on disability have been elaborated in a categorical distribution assuming a Barthel score &gt; 50 either on mobility or dependency. The results of antibody titers have been expressed as the logarithm of each value. The Stata 15.0 statistical package (Stata Corporation, College Station, TX) was used to perform all analyses. A p value of < 0.05 was considered significant.

#### Ethical approval

This study was approved by the Ethical Committee of Azienda Socio Sanitaria ULSS 8 (Del.694 del 27.05.20).

## Results

The CF, where it was conducted the epidemiological investigation and prospective data collection, is professionally staffed with 5 nurses, 20 nursing aides, 1 social worker, 1 dietician, 1 physiotherapist, etc. It is served by nursing staff 24-hour per day and an in house doctor, on demand and on fixed days, is always in attendance.

As of March 19, 2020, 64 residents (51 females and 13 males) were housed in the SCF. Forty-three out of the total 53 COVID-19 positive residents were females (81.2%) and 10 males (18.8%) with a median age of 86 (IRQ 81-92). The distribution of residents among the age groups was: 9 residents in the 65-74 years old group (16.9%), 14 in the 75-84 years old group (26.4%) and 30 subjects in the 85-99 years old group (56.6%). A total of 20 (37%) had a diagnosis of a psychiatric disorder or were affected by dementia. Major comorbidities affecting the residents enrolled were: hypertension (49%); cardiovascular diseases (45%); chronic kidney disease (41%); diabetes (26%); encephalopathy (11%). No statistical differences were found between the age groups on comorbidities and gender. Figure 1 represents the point in time prevalence of the outbreak. It shows the declining number of COVID-19 positives as they recover the rising number of deceased individuals and the stable number of always-negative subjects. From March19 to April 20, 11 COVID-19 positives residents died. The CFR was 22%. The deceased residents have been classified at severe stage of illness with respiratory symptoms. One patient with a severe stage of disease survived. The median time lag from symptoms onset and death was 7.5 days (IRQ 2.5 – 13.5) and the median age of deceased residents was 92 years old (range 79-99). The probability of mortality was 0% in the age group 65-74, 7.1% in the age group 75-85 group and 30.0% in the group of 85-99. Comorbidities were not found to be independent risk factors associated to death. A significant difference was found between the age groups in the prevalence of chronic renal diseases (p-value= < 0.04). According to the disease staging, 26 (49%) were asymptomatic, 9 (17%) had a mild disease; 7 (13%) a moderate stage and 11 (21%) a severe illness severity. A significant difference was found in the illness severity among the age groups. The 89% of subjects in the 65-75 years-old group have been asymptomatic versus 41% in the over 75 age groups (p-value= < 0.01).

**Figure 1:**
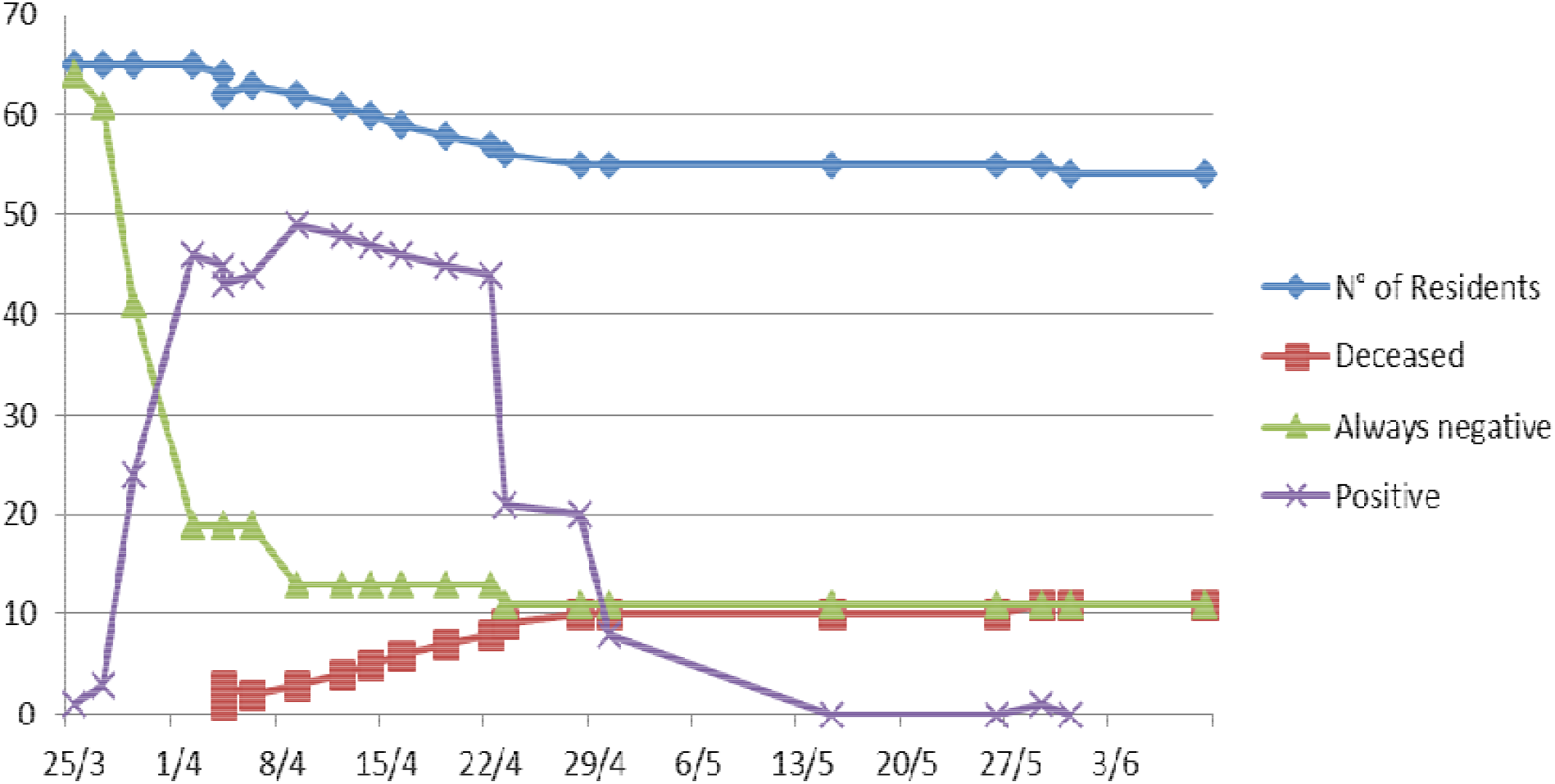
Point in time prevalence of main outcomes of the outbreak.

Table 2 analyzes the association among the main risk factors as attack rate, recovery time in the clearance of viral RNA or death. A positive age gradient was found among the age groups: as age increases, so it raises the susceptibility to contagious (p-value = 0.03) and grows the probability of death (p-value =0.05) and time to negative conversion is prolonged (p-value =0.04). Disability was positively associated to susceptibility and contagious (p value = 0,03) but not to death (p value = 0.57).

**Table 1:**
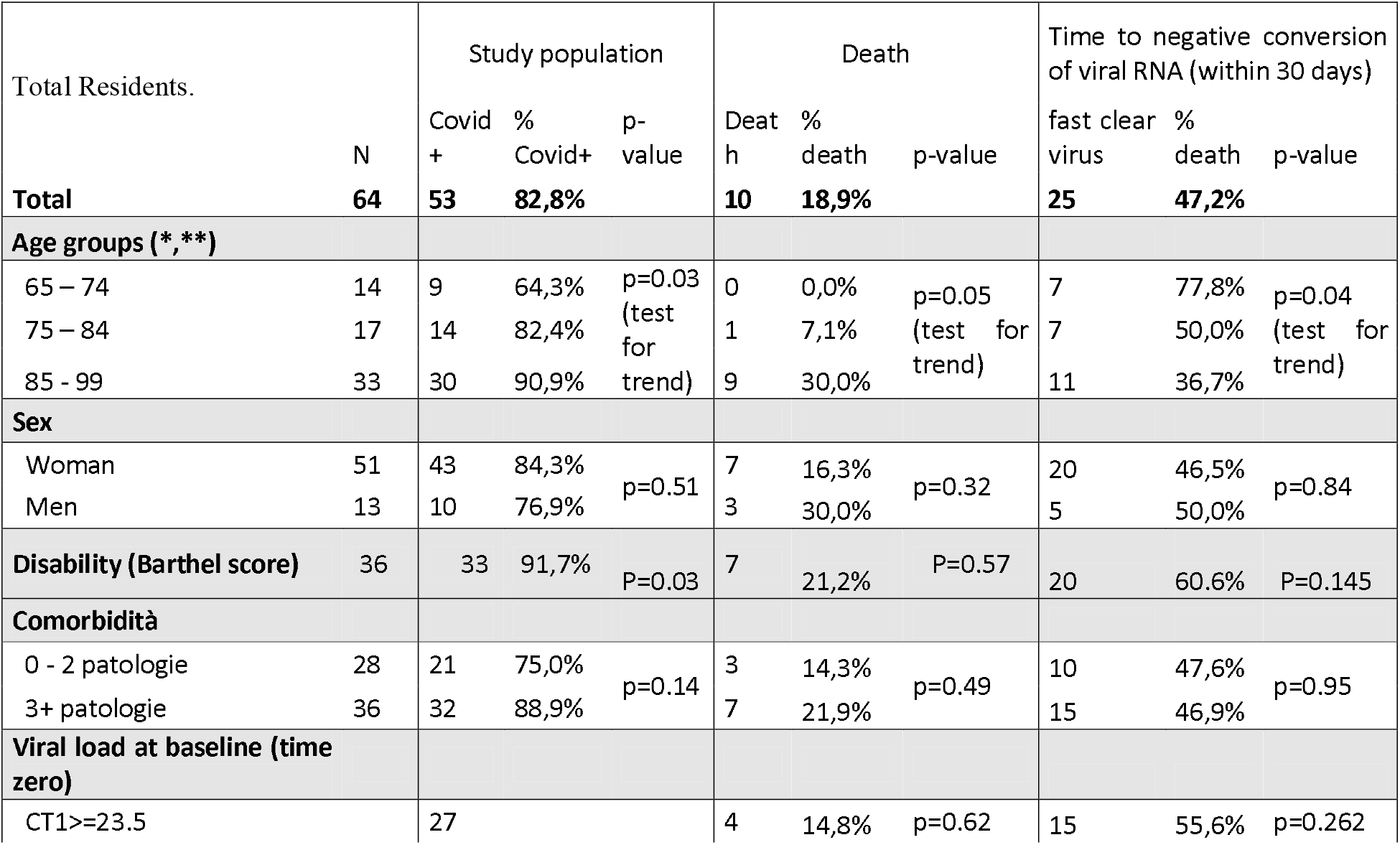
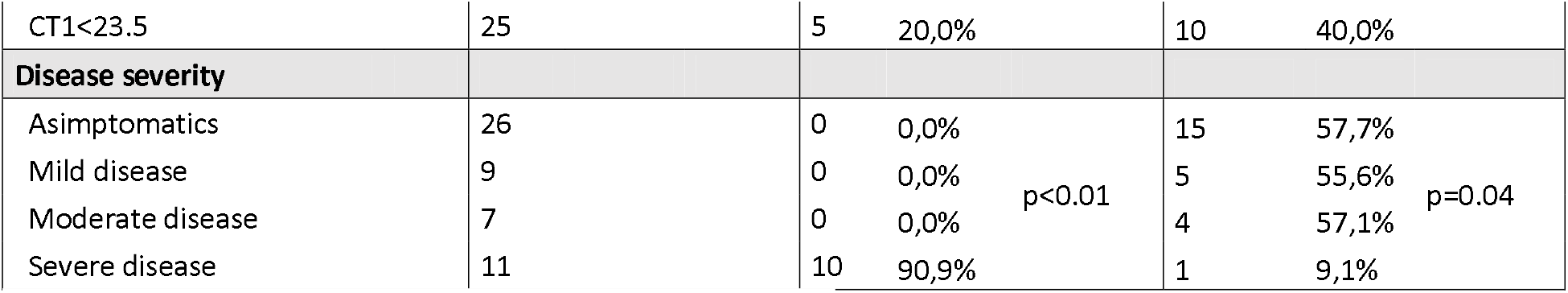
main outcomes of the epidemiological investigation.

**Table 2:** results of the retrospective study on mortality.

| Facility | Total residents<br>01/03/2020 | Covid-2<br>positives | Deceased | Attact rate | Fatality rate | SMR |
| --- | --- | --- | --- | --- | --- | --- |
| CF* | 64 | 52 | 10 | 82.8% | 22.4% | **1.11** |
| Facility 2 | 55 | 4 | 1 | 7.3% | 25.0% | **1.12** |
| Facility 3 | 121 | 47 | 11 | 38.8% | 23.4% | **1.75** |
| Facility 4 | 59 | 1 | 0 | 1.7% | 0.0% | **2.12** |
| Facility 5 | 33 | 13 | 4 | 39.4% | 30.8% | **1.01** |
| Facility 6 | 45 | 13 | 1 | 28.9% | 7.7% | **0.51** |
| Facility 7 | 78 | 42 | 9 | 53.8% | 21.4% | **3.52** |
| Facility 8 | 146 | 23 | 11 | 15.8% | 47.8% | **1.57** |
| Facility 9 | 85 | 70 | 13 | 82.4% | 18.6% | **2.17** |
| **Total** | **686** | **262** | **61** | **38.2%** | **23.3%** | **1,6** |

\*CF: Case Facility

No association was found between viral load at baseline, gender and comorbility. Younger residents had a higher viral load. (77.8% in the 65-75 years old group had Ct<23.5 vs. 43% in the age group of over 85 years-old). In spite of the higher viral load, the subjects of youngest group were significantly more asymptomatic than those in the oldest age groups: 88% in the 65-74 years old group; 50% in the 75-84 years old; 36% in the over 85 (p-value=0, 02). Twenty-two residents converted to negativity RNA virus within the third and 13 in the fourth week. A remaining group of 8 residents cleared the virus at the sixth week. The latest resident cleared in 50 days. It is also worth to note that 2 residents had an alternate result in nasopharyngeal swab sample as two negative or indeterminate results were followed by a positive one.

Immune response was appraised measuring antibodies at the fourth and after 16 weeks after the date of infection. In figure 2, it is shown that the oldest groups had a high titer antibody response that was maintained over time.

**Figure 2:**
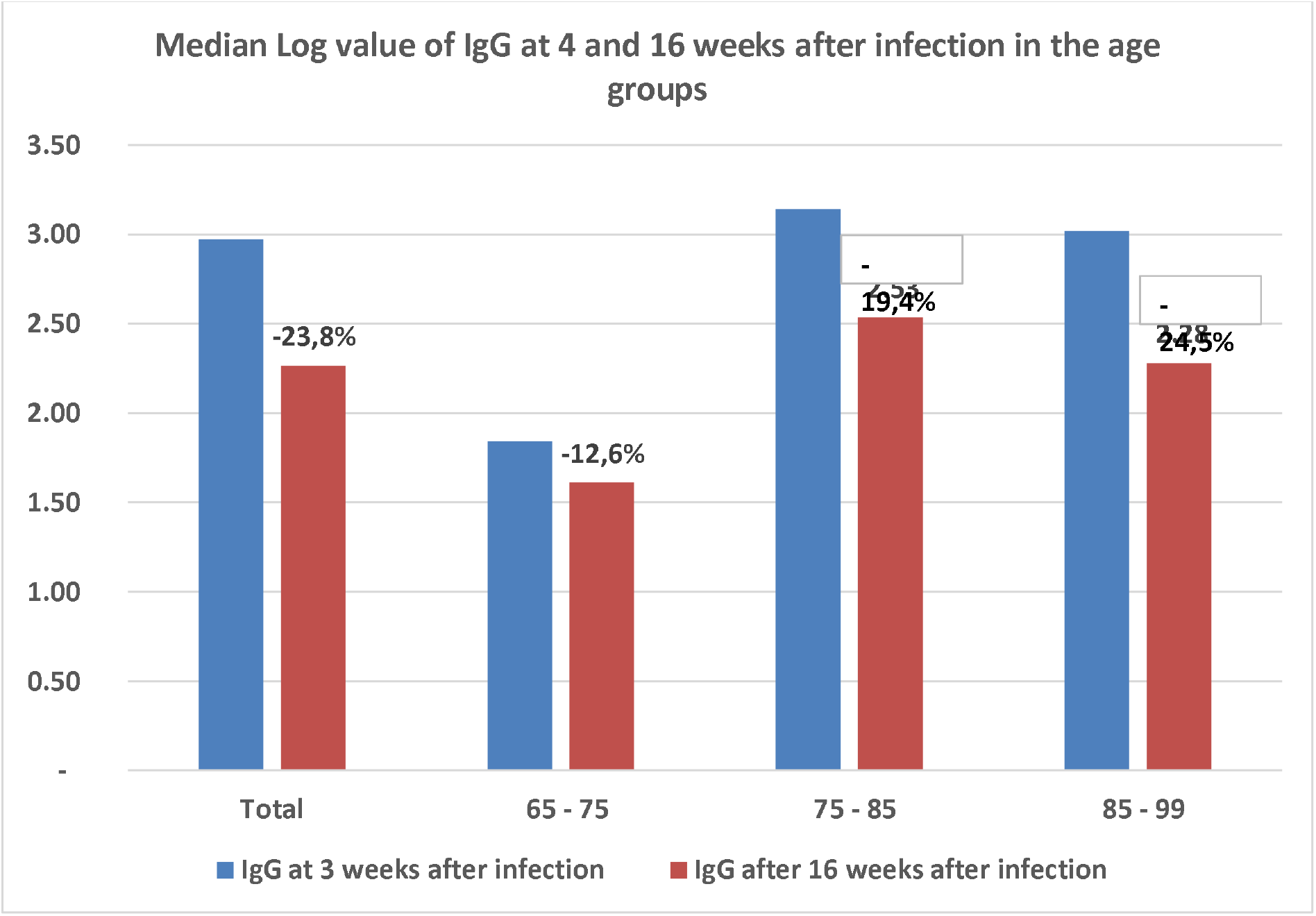
The antibody titer after 4 and 16 weeks.

The retrospective analysis was conducted in other 8 facilities out of 34 of the Vicenza Province, which reported 262 COVID-19 cases. In total, from February 20 to June 15, the number of residents hosted in these facilities was 686 with a median age of 88 years-old (IQ 82-92-81% female, 19% male). Table 3 shows the different impact of the epidemics in these facilities. The attack rate ranged from 7.3% to 82.8%. Sixty-one out of the 262 residents, tested positives, deceased. The CFR also widely ranged from 7.7% to 47.8%.

The median age of deceased residents was 90 years-old (IQ 85-94 - 80% female, 20% male). The SMR demonstrated a total increase of deaths of 60% if compared with those of 2018 and 2019. In the period considered, 150 deaths were observed toward a number of 93 expected. It should be noted that in the Facility 4, only 1 deceased individual was tested positive against a SMR of 2.12%. This leads to the conclusion that these figures may underestimate the real impact on mortality. Outbreak control measures

## Discussion

Residents of LTCFs are old and frail, with complex health needs and underlying comorbidities. LTCF, or other care setting with different definition, have in common to be environments where a relatively large number of people congregate, and are consequently prone to infection diseases outbreaks. The fast spread of respiratory virus outbreaks, including flu, in LTCFs is well recognized [10]. Besides the COVID-19 pandemic, LTCFs are also a reservoir of bacteria resistant to antimicrobials. Patients resident in LTCFs are often extensively colonized with potential pathogens such as Staphylococcus aureus, beta-hemolytic streptococci, members of the Enterobacteriaceae, or Pseudomonas aeruginosa [11].

Diagnosis in LTCFs is a difficult matter. Hearing and cognition are often impaired in LTCF residents. Symptoms may not be expressed or correctly interpreted by caregivers. Many single reports highlighted the vulnerability of elderly to COVID-19 and it first became evident after the first outbreak in a skilled nursing facility within King County, Washington, late February 2020 and in other settings [12, 13, 14]. The attributable cause of deaths by COVID-19 in LTCFs is one of major limit reported in the literature. Some countries recorded data without a test and with a diagnosis based only to clinical symptoms. The place of death is another possible bias as many deaths were reported in hospital instead of LTCFs [5]. Under-ascertainment and under reporting of COVID-19 cases has been a common feature of the pandemic.

International evidence of mortality associated with COVID-19 in LTCFs has been published and periodically updated. The impact of COVID-19 on LTCFs residents has been very different internationally. However, using data with the caveats that definitions used and difficulties in comparing data, it has been estimated that the share of total COVID-19 associated deaths in residents of LTCFs is 47% (based on 26 countries) [15]. In Ontario, Canada, the population in LTCFs represented over 80% of deaths from probable cases of COVID-19 [16]. A report based on different sources of data and from different regions of the world estimated a CFR of 14.8% in the age class > 80. [17].

The SMR calculated in the subset of LTCFs of the Vicenza Province is up to 60%. The comparison of CFR and age of death between the group of 8 LTCF and the CSF were similar. The median age of deceased residents was 90 years old in the 9 LTCF and 92 years old in the SCF, the CFR was 23.3% while in SCF was equal to 22.4%. We found age gradient in all dimensions considered (i.e. symptoms, illness severity, time of recovery, clearance of the virus, immune response and CFR). We did not find a positive correlation of comorbidity with death and it may be due to the small sample of the population studied. A meta-analysis evidenced that hypertension, diabetes, chronic obstructive pulmonary disease (COPD), cardiovascular disease and cerebrovascular disease are major risk factors associate with Covid-19 adverse outcome. Liver disease, malignancy or renal disease had no correlation with death [19]. In our population, over 40% of residents were affected by a chronic renal disease. This unusual frequency of such disease in our study population may in part justify the lack of association. Data on serology, particularly those obtained 16 weeks after contagious, demonstrated that many individuals had a strong immune response. The association between mortality and age together with age and a high antibody titer would lead to consider that it could be the mark of an unbalanced and ineffective immune response. Pathophysiology of Covid-19 might involve the immune system and its age associated alterations, known as immunosenescence, may produce a progressive inability to respond to infections.

Our work has the many limits. First is the poor statistical power which does not permit to generalize any conclusion of the observational study. Moreover, in the calculation of SMR we were not able to report on deceased individuals without a COVID-19 test. The diagnosis of COVID-19 associated death was made on a clinical basis and it might lead to diagnostic mistakes. What might have a certainty is that COVID-19 highlighted the neglect of quality improvements in these settings of care and the need for adoption of medical standards by the European countries [21].

## Data Availability

The dataset is not available on external repositories

